# Resting heart rate as a biomarker for tracking change in cardiorespiratory fitness of UK adults: The Fenland Study

**DOI:** 10.1101/2020.07.01.20144154

**Authors:** Tomas I. Gonzales, Justin Y. Jeon, Timothy Lindsay, Kate Westgate, Ignacio Perez-Pozuelo, Stefanie Hollidge, Katrien Wijndaele, Kirsten Rennie, Nita Forouhi, Simon Griffin, Nick Wareham, Soren Brage

## Abstract

**Aims:** Resting heart rate (RHR) is inversely associated with cardiorespiratory fitness (CRF) but few studies have investigated the nature of this relationship in large population samples. We examined the association between RHR and CRF in UK adults and explored factors that may influence this relationship.

**Methods and Results:** In a population-based sample of 5,143 men and 5,722 women (aged 29-65 years), mean (SD) RHR while seated, supine, and during sleep was 67.6 (9.8), 63.5 (8.9), and 56.9 (6.9) bpm, respectively. The age- and sex-adjusted association with CRF as assessed by submaximal treadmill testing was −0.26 (95%CI −0.27; −0.24), −0.31 (95%CI −0.33; −0.29), and −0.31 (95%CI −0.34; −0.29) ml O_2_ kg^-1^ beat^-1^. Sequential adjustment for objectively measured obesity and physical activity attenuated the RHR coefficient by 10% and 50%, respectively. In longitudinal analyses of 6,589 participants re-examined after 6 years, each 1 bpm increase in supine RHR was associated with 0.23 (95%CI 0.20; 0.25) ml O_2_ min^-1^ kg^-1^ decrease in CRF.

**Conclusions:** Across all measures, RHR is inversely associated with CRF; half of this association is explained by obesity and physical activity, suggesting CRF changes achieved through altered behaviour could be tracked through changes in RHR, a notion supported by longitudinal results. As well as its utility as a biomarker of CRF at population-level, serial measurements of RHR may facilitate personal goal setting/evaluation and remote patient monitoring.

## Introduction

Cardiorespiratory fitness (CRF) is associated with the incidence of type 2 diabetes, cardiovascular disease, various types of cancer (1–3), as well as all-cause and cause-specific mortality (4,5). In spite of strong evidence of its association with clinical outcomes, it is relatively uncommon for CRF to be measured in clinical care, mostly because its assessment using graded exercise testing is time-consuming, costly and potentially unsafe for some patient groups without medical supervision. Measurement of resting heart rate (RHR) could serve as a viable alternative to clinical CRF testing as it is easy to undertake and is scalable to large populations through the use of wearable sensors. It also has a pattern of association with future health endpoints that is similar to that of CRF (6–12).

The potential clinical utility of using RHR as a proxy for CRF has not been realised in part due to unresolved methodological challenges. RHR values are known to be dependent on the physiological state at the time of measurement (13). In clinical practice, the most common states in which RHR measurement is taken are sitting during blood pressure measurement or lying supine during brief multi-lead ECG measurement. In free-living individuals, however, wearable sensors offer the potential for convenient assessment of heart rate in other states of rest, particularly sleep, and arousal. It is unknown whether differences in RHR measured in these different ways alters its relationship with CRF. It is also unknown to what extent the relationship between RHR and CRF is affected by factors such as adiposity and physical activity, which have established impact on CRF (14,15). Quantifying the degree to which these modifiable factors may explain the relationship between RHR and CRF would strengthen the argument for using RHR as a proxy measure of exercise capacity. Finally, although some studies have described the longitudinal relationship between RHR and CRF (8,16), there is uncertainty about how individual changes in RHR may reflect longer-term changes in CRF in the general population.

Here, we aimed to assess the cross-sectional associations between different measures of RHR and CRF in a large population-based study of UK adults, and how body composition and physical activity may alter the relationship between RHR and CRF. Secondly, we examined whether within-person change in RHR is associated with within-person change in CRF in longitudinal analysis.

## Methods

### Participants

Participants were recruited by letter from general practice lists in and around Cambridgeshire in the East of England to the Fenland Study, a population-based cohort study designed to investigate interactions between environmental and genetic factors in determining obesity, type 2 diabetes, and related metabolic disorders in young to middle-aged adults (17). Exclusion criteria included prevalent diabetes, pregnancy or lactation, inability to walk unaided, psychosis or terminal illness (life expectancy of ≤ 1 year at the time of recruitment). The study complies with the Declaration of Helsinki and was approved by the Health Research Authority NRES Committee East of England-Cambridge Central. All participants gave written informed consent.

Participants were invited to attend one of three testing facilities (Ely, Wisbech or Cambridge). Recruitment from general practice surgeries began in 2005, collecting data on a total of 12,435 participants (response rate 27%) as previously described (17). For the present analysis, data from 10,865 individuals was included after excluding participants with key missing variables such as RHR, VO_2_max, physical activity and body composition, as well as participants prescribed beta-blockers (n=315). Compared to the main cohort sample, included participants were 0.4 years younger, 2% more physically active and had a 0.2 bpm lower RHR. They were also 1 cm taller, had 0.5 kg lower body mass and 0.5% less bodyfat but were otherwise similar across other demographic characteristics (Supplementary Table 1).

**Table 1.**
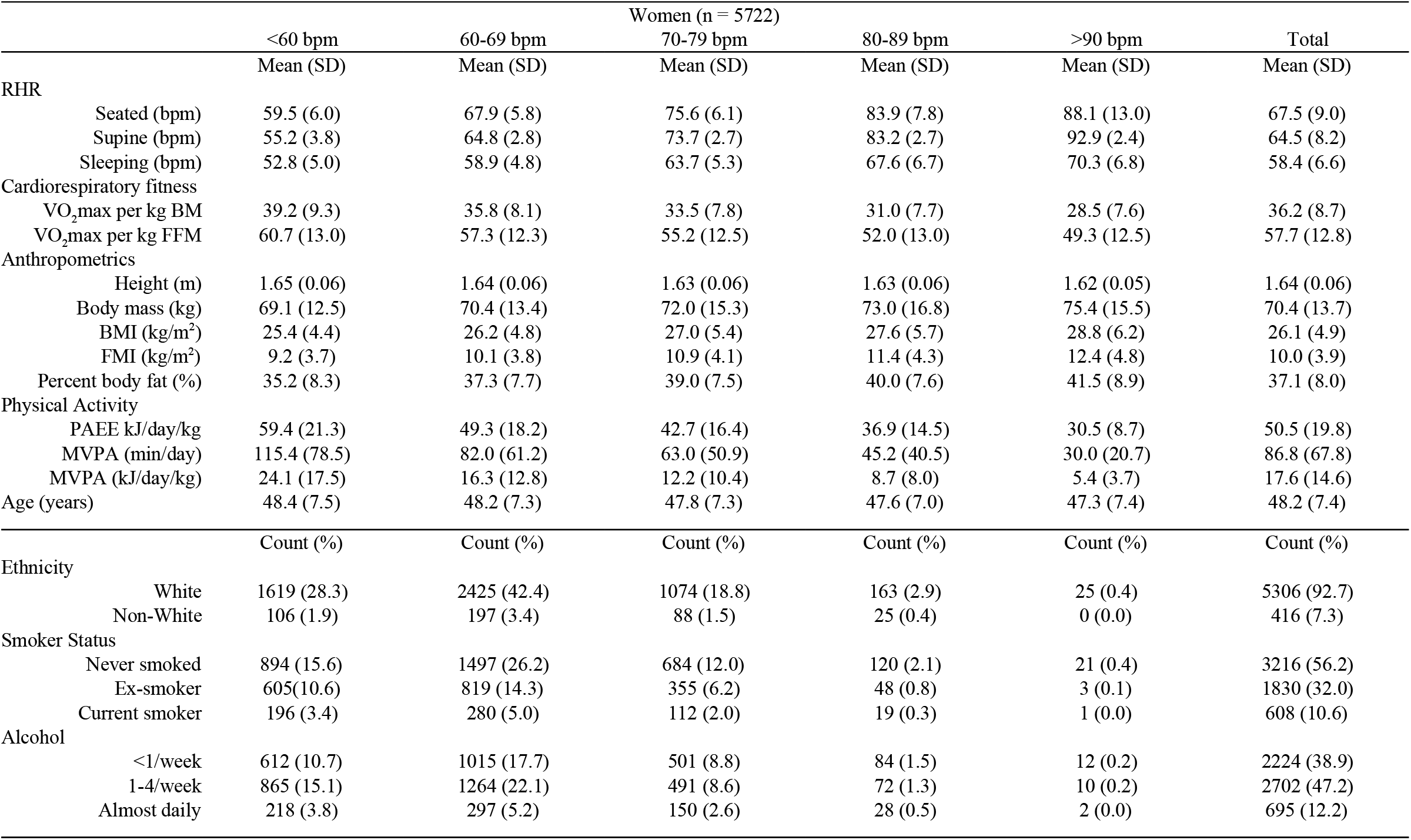
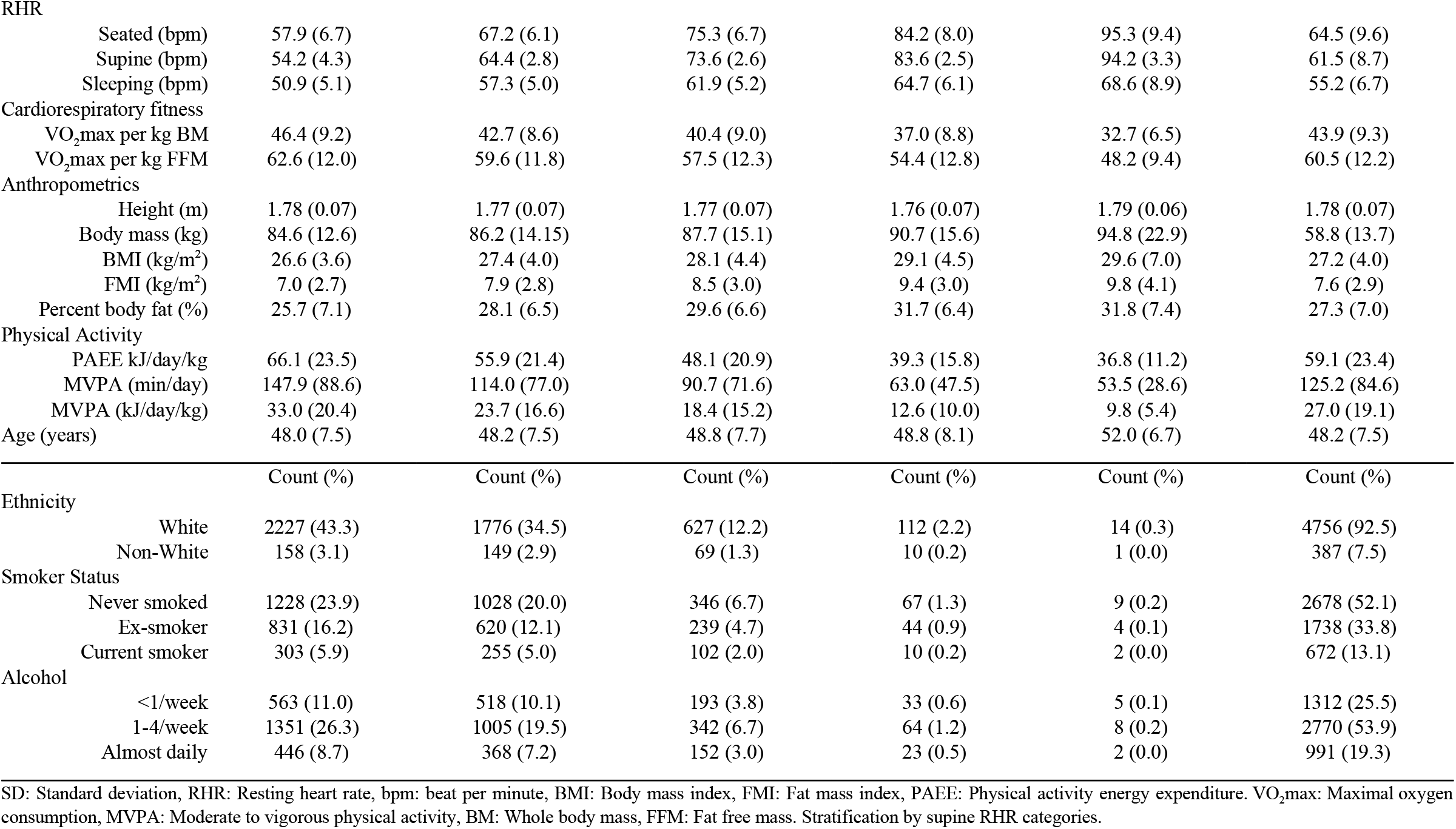
Baseline participant characteristics in women and men. The Fenland Study.

RHR and CRF were assessed again after a median (interquartile range) of 6 (5-8) years in data that was available at the time of analysis from a subsample of 6,589 participants allowing the examination of the relationship between within-person change in RHR and CRF over time.

### Experimental procedures

Participants arrived at the testing facility after an overnight fast to complete baseline clinical assessment and questionnaires. At least half an hour after arrival, resting pulse rate was measured in a seated position while resting blood pressure was being assessed using an Omron 705CP-II (OMRON Healthcare Europe, Hoofddorp, Netherlands) three times, at 1 minute intervals. The mean of these three pulse rate values was used as seated RHR. At least one hour after arrival, RHR was measured with the participant at rest in a supine position using a combined heart rate (HR) and movement sensor (Actiheart, CamNtech, Papworth, UK) attached to the chest at the base of the sternum by two standard ECG electrodes (18,19). HR was recorded for 6 minutes and RHR was calculated as the mean HR measured during the last 3 minutes. Following the visit to the testing facility, participants wore the combined HR and movement sensor continuously for 6 days and nights during free-living, recording at 60-second intervals. The daily minimum HR, determined robustly as the thirtieth lowest minute-by-minute HR reading during each 24-hour period, was averaged across the days to estimate habitual sleeping HR (20).

VO_2_max was assessed using HR response to a submaximal treadmill test. Participants exercised while treadmill speed and grade were progressively increased across several stages of level walking, inclined walking, and level running. The test was terminated if one of the following criteria were met: 1) the participant wanted to stop, 2) the participant reached 90% of age-predicted maximal HR (208 − 0.7 · age) (21); or 3) the participant had exercised above 80% of age-predicted maximal HR for >2 minutes. Predicted workload (physical activity intensity) during the treadmill protocol (20) was regressed against HR (expressed in bpm above sleeping HR) to define the individual’s submaximal relationship between HR and physical activity intensity. The HR-to-activity intensity relationship was then extrapolated to age-predicted maximal HR to predict maximal work capacity. The resulting work capacity was converted to VO_2_max by adding an estimate of resting metabolic rate (RMR) (22) and dividing by the energetic equivalent of oxygen (23). VO_2_max was expressed in both ml O_2_ min^-1^ kg^-1^ body mass and as ml O_2_ min^-1^ kg^-1^ fat-free mass. The VO_2_max estimation procedure has been validated against directly measured VO_2_max, demonstrating low bias and high correlation (24).

Ethnicity (White, non-White), smoking status (never, former, current), and alcohol intake (units/week) were determined using a self-administered questionnaire. Anthropometric measures were collected by trained personnel. Weight was measured with a calibrated electronic scale (TANITA model BC-418 MA; Tanita, Tokyo, Japan) and height was assessed with a wall-mounted, calibrated stadiometer (SECA 240; Seca, Birmingham, United Kingdom). Body composition was assessed using dual-energy X-ray absorptiometry (DEXA) (GE Lunar Prodigy Advanced fan beam scanner, GE Healthcare, Bedford, United Kingdom) deriving fat, lean and bone mass measurements across all body regions (25).

Physical activity was objectively measured over 6 days using a combined HR and movement sensor (Actiheart, CamNtech, Papworth, UK) with individual calibration of the HR-to-physical activity intensity relationship performed using data from the treadmill test as described above (17). Free-living data was pre-processed (26) and modelled using a branched equation framework (27) to estimate intensity time-series, which was then summarised over time as daily physical activity energy expenditure (PAEE) (kJ/kg per day) (17). Intensity was also expressed in standard metabolic equivalents (METs), using the conversion 1 MET = 71 J/min/kg (∼3.5 ml O_2_ min^-1^ kg^-1^), and summarised as moderate-to-vigorous physical activity for intensity greater than 3.0 METs. In the present analyses, the proportional PAEE accumulated in moderate and vigorous physical activity was used alongside total PAEE to account for both total physical activity and intensity in the same model (17).

For the longitudinal analyses in the subsample, supine RHR was measured again at follow-up using a 15-min rest protocol but the mean of minutes 4 to 6 was used in the present analysis to match the baseline design for derivation of change in supine RHR. CRF was assessed in the same way as baseline.

### Statistical analyses

We used sex-stratified regression models to examine cross-sectional associations between RHR and CRF while adjusting for groups of confounding or explanatory factors. The first model (model 1) only adjusted for age, whilst model 2 additionally adjusted for demographic and lifestyle factors (ethnicity, smoking status, alcohol intake). The third model (model 3) additionally adjusted for BMI. The fourth model (model 4) added adjustment for PAEE, and the fifth model (model 5) further adjusted for moderate to vigorous intensity activity (as a fraction of total PAEE). When cardiorespiratory fitness was expressed relative to fat-free mass (FFM) instead of body mass, fat mass index (fat mass divided by height squared) was used as the adjustment for adiposity instead of BMI. Subgroup analyses were performed as follows: Analyses were stratified by groups of age (less than 50y; 50-59y; 60y and greater), BMI (normal weight, BMI 18.5-25; overweight, BMI 25-30; obese, BMI above 30 kg·m^-2^), and PAEE level (<40, 40-60, >60 kJ·kg^-1^ day^-1^). Descriptive statistics were reported as means and standard deviations, unless specified otherwise. Interrelationships of RHR measures were examined by simple regression. For the longitudinal analysis of the subsample with repeat measures of RHR and CRF, the association between within-person change in RHR and CRF was adjusted for baseline age, sex, RHR, and fitness, as well as age at follow-up. Binscatter plots (5% bins) were used to visualise associations between RHR and VO_2_max. Statistical analyses were performed with STATA (Version 14.2; StataCorp, College Station, TX); a p-value of 0.05 or less was considered statistically significant for all analyses.

## Results

Mean (SD) baseline RHR in the seated position, supine position, and during sleep was 67.6 (9.8), 63.5 (8.9), and 56.9 (6.9) bpm, respectively (Figure 1), and correlations (Pearson r) between these modalities ranged from 0.65 to 0.81 (Supplementary Table 2). On average, RHR was 3 bpm higher and VO_2_max was 7.7 ml O_2_ min^-1^ kg^-1^ body mass lower in women compared to men. Characteristics of participants included for analyses are summarised in Table 1, stratified by sex and supine RHR categories. Those with higher RHR had higher BMI and body fat levels, lower VO_2_max values, and were less physically active.

**Figure 1.**
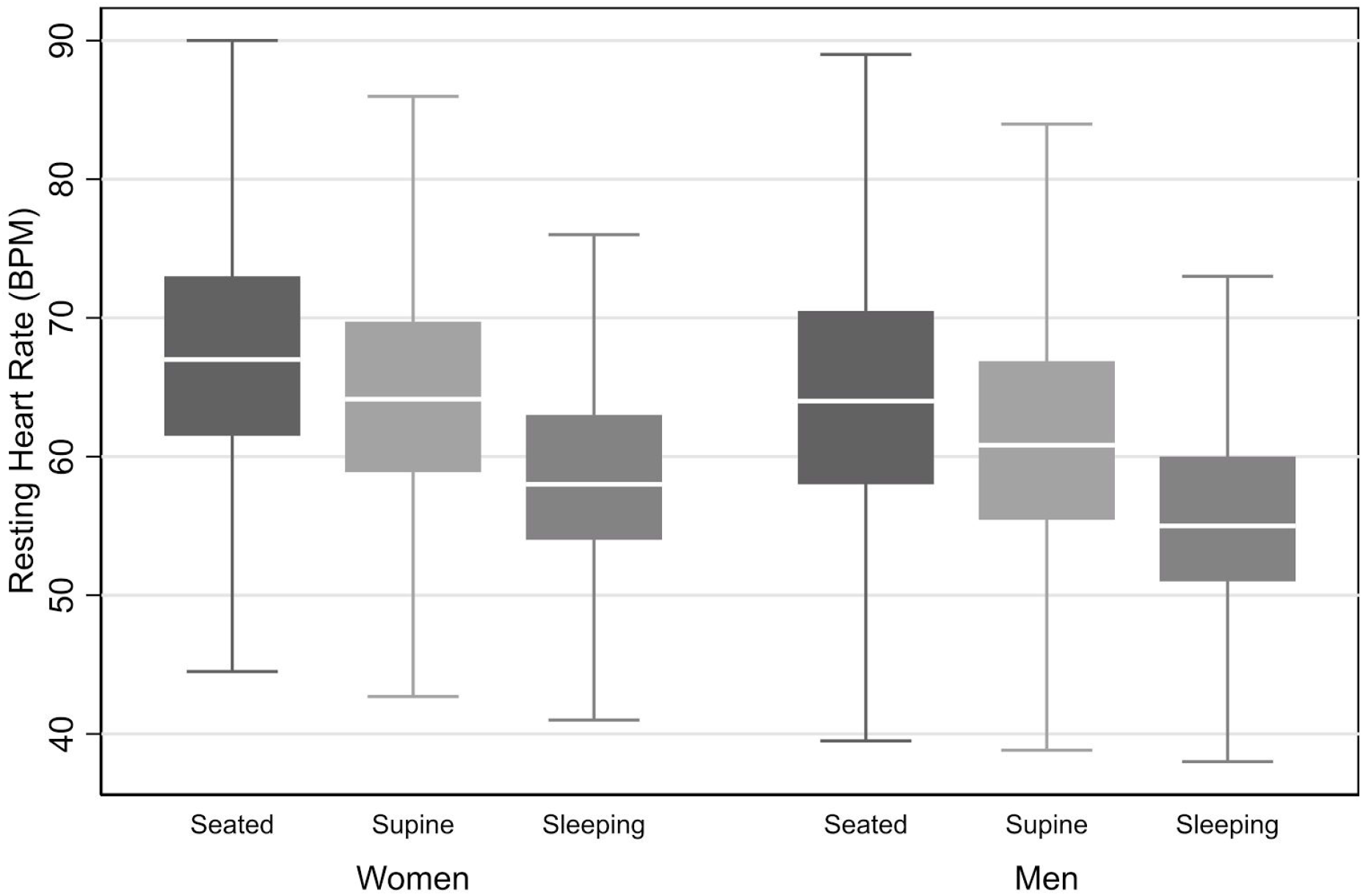
Resting heart rate stratified by sex and measurement modality. The Fenland Study (n=10,865)

**Table 2.** Association between resting heart rate and maximal oxygen consumption expressed per kg whole-body mass. The Fenland Study

|  |  | Seated RHR |  | Supine RHR |  | Sleeping RHR |  |
| --- | --- | --- | --- | --- | --- | --- | --- |
|  |  | Men | Women | Men | Women | Men | Women |
| Total sample |  |  |  |  |  |  |  |
| Model 1 |  | -0.27 (-0.29, -0.24) | -0.25 (-0.27, -0.22) | -0.33 (-0.36, -0.30) | -0.31 (-0.33, -0.28) | -0.28 (-0.32, -0.24) | -0.27 (-0.31, -0.24) |
| Model 2 |  | -0.27 (-0.29, -0.24) | -0.25 (-0.27, -0.22) | -0.32 (-0.35, -0.29) | -0.30 (-0.33, -0.28) | -0.31 (-0.35, -0.28) | -0.31 (-0.34, -0.28) |
| Model 3 |  | -0.23 (-0.26, -0.21) | -0.19 (-0.22, -0.17) | -0.29 (-0.32, -0.26) | -0.26 (-0.28, -0.23) | -0.26 (-0.30, -0.23) | -0.22 (-0.25, -0.19) |
| Model 4 |  | -0.16 (-0.18, -0.14) | -0.13 (-0.15, -0.11) | -0.18 (-0.21, -0.16) | -0.17 (-0.19, -0.14) | -0.17 (-0.21, -0.13) | -0.14 (-0.17, -0.11) |
| Model 5 |  | -0.14 (-0.16, -0.12) | -0.12 (-0.14, -0.10) | -0.16 (-0.18, -0.13) | -0.15 (-0.17, -0.12) | -0.15 (-0.18, -0.11) | -0.13 (-0.16, -0.10) |
| Age stratified |  |  |  |  |  |  |  |
| Model 2 | <50 years | -0.26 (-0.30, -0.22) | -0.25 (-0.29, -0.21) | -0.34 (-0.38, -0.29) | -0.29 (-0.34, -0.25) | -0.35 (-0.41, -0.29) | -0.30 (-0.35, -0.24) |
|  | 50-60 years | -0.26 (-0.30, -0.23) | -0.25 (-0.28, -0.21) | -0.30 (-0.35, -0.26) | -0.31 (-0.35, -0.27) | -0.28 (-0.34, -0.22) | -0.32 (-0.37, -0.27) |
|  | >60 years | -0.28 (-0.33, -0.22) | -0.24 (-0.30, -0.19) | -0.33 (-0.39, -0.27) | -0.32 (-0.38, -0.26) | -0.31 (-0.40, -0.23) | -0.33 (-0.41, -0.25) |
| Model 3 | <50 years | -0.22 (-0.26, -0.18) | -0.20 (-0.23, -0.16) | -0.30 (-0.34, -0.26) | -0.25 (-0.29, -0.21) | -0.29 (-0.35, -0.23) | -0.21 (-0.26, -0.16) |
|  | 50-60 years | -0.23 (-0.27, -0.19) | -0.19 (-0.22, -0.15) | -0.27 (-0.31, -0.23) | -0.25 (-0.29, -0.22) | -0.23 (-0.29, -0.17) | -0.22 (-0.27, -0.17) |
|  | >60 years | -0.25 (-0.31, -0.19) | -0.21 (-0.27, -0.16) | -0.30 (-0.36, -0.24) | -0.29 (-0.35, -0.23) | -0.28 (-0.36, -0.20) | -0.27 (-0.34, -0.19) |
| Model 5 | <50 years | -0.12 (-0.16, -0.08) | -0.10 (-0.14, -0.07) | -0.15 (-0.19, -0.11) | -0.11 (-0.15, -0.07) | -0.14 (-0.20, -0.09) | -0.10 (-0.15, -0.05) |
|  | 50-60 years | -0.15 (-0.18, -0.11) | -0.11 (-0.14, -0.08) | -0.15 (-0.19, -0.11) | -0.14 (-0.18, -0.10) | -0.13 (-0.19, -0.08) | -0.12 (-0.17, -0.07) |
|  | >60 years | -0.16 (-0.21, -0.11) | -0.17 (-0.22, -0.12) | -0.17 (-0.24, -0.11) | -0.22 (-0.28, -0.16) | -0.17 (-0.25, -0.10) | -0.22 (-0.29, -0.15) |
| BMI stratified |  |  |  |  |  |  |  |
| Model 3 | <25 kg/m<sup>2</sup> | -0.30 (-0.35, -0.26) | -0.24 (-0.27, -0.20) | -0.35 (-0.40, -0.30) | -0.32 (-0.36, -0.28) | -0.34 (-0.41, -0.27) | -0.31 (-0.36, -0.26) |
|  | 25-30 kg/m<sup>2</sup> | -0.21 (-0.24, -0.17) | -0.18 (-0.22, -0.14) | -0.27 (-0.31, -0.24) | -0.22 (-0.26, -0.17) | -0.24 (-0.30, -0.19) | -0.17 (-0.22, -0.11) |
|  | >30 kg/m<sup>2</sup> | -0.18 (-0.23, -0.13) | -0.11 (-0.15, -0.06) | -0.22 (-0.28, -0.17) | -0.17 (-0.21, -0.12) | -0.19 (-0.26, -0.11) | -0.13 (-0.19, -0.07) |
| Model 5 | <25 kg/m<sup>2</sup> | -0.18 (-0.23, -0.14) | -0.13 (-0.17, -0.10) | -0.18 (-0.23, -0.13) | -0.17 (-0.21, -0.14) | -0.18 (-0.24, -0.11) | -0.18 (-0.23, -0.14) |
|  | 25-30 kg/m<sup>2</sup> | -0.13 (-0.16, -0.09) | -0.13 (-0.17, -0.09) | -0.15 (-0.19, -0.12) | -0.13 (-0.17, -0.08) | -0.15 (-0.20, -0.10) | -0.09 (-0.14, -0.04) |
|  | >30 kg/m<sup>2</sup> | -0.10 (-0.15, -0.05) | -0.08 (-0.12, -0.04) | -0.12 (-0.17, -0.06) | -0.11 (-0.16, -0.07) | -0.08 (-0.16, -0.01) | -0.08 (-0.14, -0.02) |
| PAEE stratified |  |  |  |  |  |  |  |
| Model 5 | <40 kJ/day/kg | -0.13 (-0.18, -0.08) | -0.14 (-0.18, -0.11) | -0.18 (-0.23, -0.13) | -0.18 (-0.22, -0.14) | -0.11 (-0.19, -0.04) | -0.13 (-0.18, -0.08) |
|  | 40-60 kJ/day/kg | -0.15 (-0.19, -0.11) | -0.10 (-0.13, -0.06) | -0.17 (-0.22, -0.13) | -0.12 (-0.16, -0.08) | -0.16 (-0.21, -0.10) | -0.13 (-0.17, -0.08) |
|  | >60 kJ/day/kg | -0.13 (-0.17, -0.09) | -0.10 (-0.15, -0.06) | -0.12 (-0.16, -0.07) | -0.12 (-0.17, -0.07) | -0.16 (-0.22, -0.11) | -0.14 (-0.20, -0.08) |

Reported values are $\beta$ coefficients (95%CI).
Model 1: Age-adjusted
Model 2: Model 1 + ethnicity, smoking and alcohol adjusted
Model 3: Model 2 + body mass index (BMI) adjusted
Model 4: Model 3 + physical activity energy expenditure (PAEE) adjusted
Model 5: Model 4 + moderate-vigorous intensity PAEE adjusted

We used several regression models to examine the association between RHR and VO_2_max per kg body mass (Table 2, Figure 2) and per kg fat-free mass (Supplementary Table 3, supplementary Figure 1). In models only adjusted for age, RHR was significantly associated with VO_2_max in women and men, irrespective of how RHR was measured. These associations remained almost identical after additional adjustment for ethnicity, smoking, and alcohol use (Model 2). Further adjustment by BMI (Model 3) attenuated the strength of associations by about 10% for VO_2_max per kg body mass. The equivalent FMI adjustment for RHR associations with VO_2_max per kg fat-free mass resulted in stronger associations, particularly for sleeping RHR for which beta coefficients were 60% larger in magnitude.

**Figure 2.**
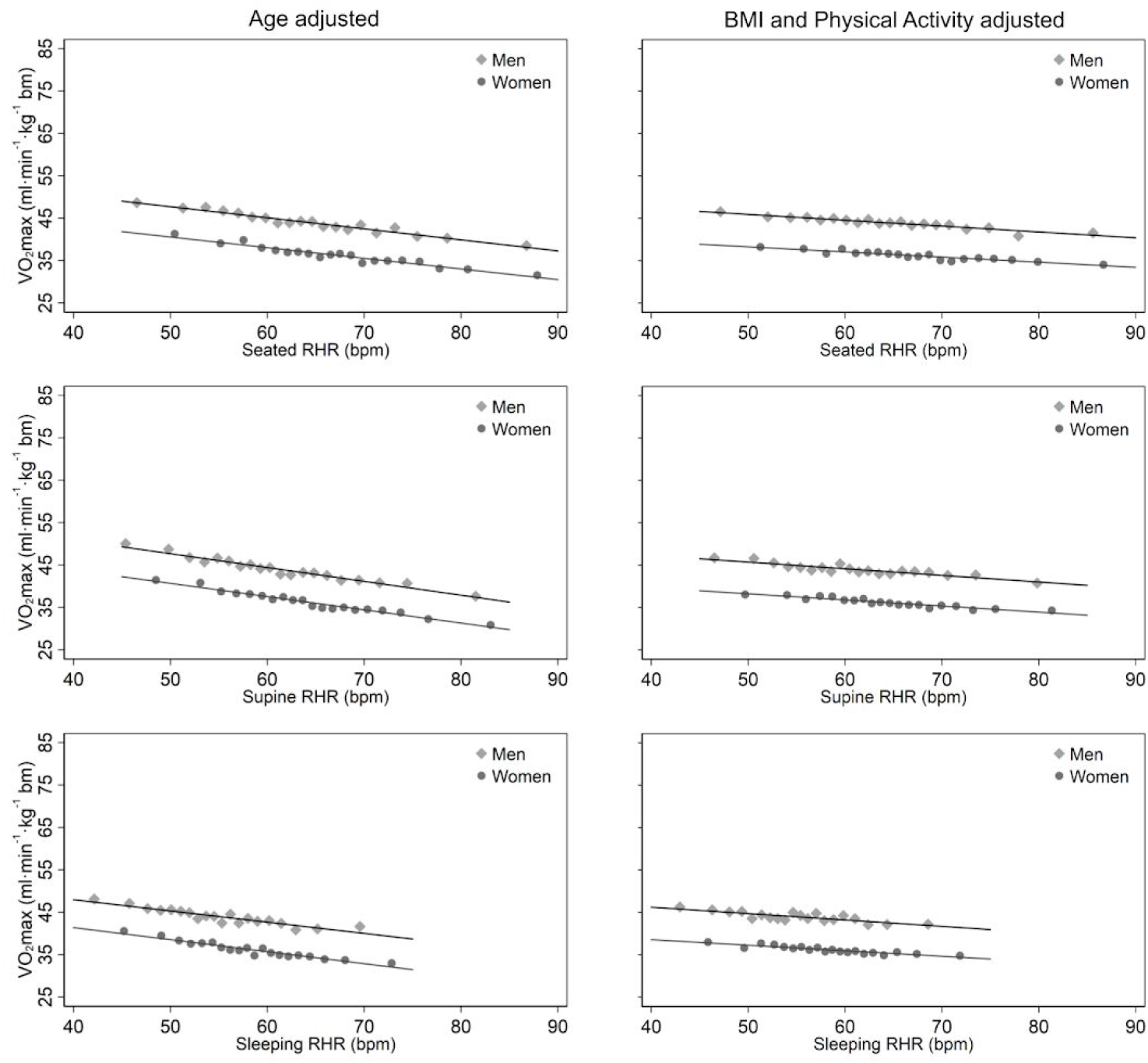
Associations between resting heart rate and maximal oxygen consumption expressed per kg body mass, stratified by sex and adjusted for age (left column panels) or age, ethnicity, smoking, alcohol consumption, body mass index, physical activity energy expenditure (PAEE), moderate-vigorous PAEE (right column panels). Top: Seated resting heart rate. Middle: Supine resting heart rate. Bottom: Sleeping resting heart rate. The Fenland Study (n=10,865). Each point represents 5% of data in the binscatter plots.

Adjustment for PAEE (Model 4) attenuated the RHR-VO_2_max relationship by 30-40% irrespective of how VO_2_max was expressed, with 5-15% additional attenuation by also accounting for the proportion of PAEE expended at moderate and vigorous intensity (Model 5). Associations between RHR and VO_2_max were relatively similar across all RHR measurement modalities, especially in the maximally adjusted models, but BMI and physical activity attenuated more of the relationship for sleeping HR in women (30% plus another 40%, respectively). We also analysed the association between RHR and VO_2_max across age, BMI, and PAEE strata separately. In general, associations in these subgroups were similar in strength to pooled associations and all remained statistically significant.

In the subsample of participants (n = 6,589) with 6-yr repeat measures of supine RHR and VO_2_max, the overall group mean levels of RHR and fitness were relatively similar to baseline values but with a wide range of individual changes over time; the 5^th^ to the 95^th^ percentiles of changes being −11.4 to 9.2 bpm and −10.1 to 10.8 ml O_2_ min^-1^ kg^-1^, respectively. In the longitudinal association analysis, each 1 bpm increase in supine RHR was associated with a 0.23 (95%CI 0.20; 0.25) ml O_2_ min^-1^ kg^-1^ decline in VO_2_max, adjusted for follow-up time and baseline values of age, sex, RHR, and VO_2_max. Sex-stratified coefficients were −0.21 (95%CI −0.24; −0.17) and −0.25 (95%CI −0.28; −0.21) ml O_2_/min/kg per 1 bpm increase in RHR in women and men, respectively (Figure 3).

**Figure 3.**
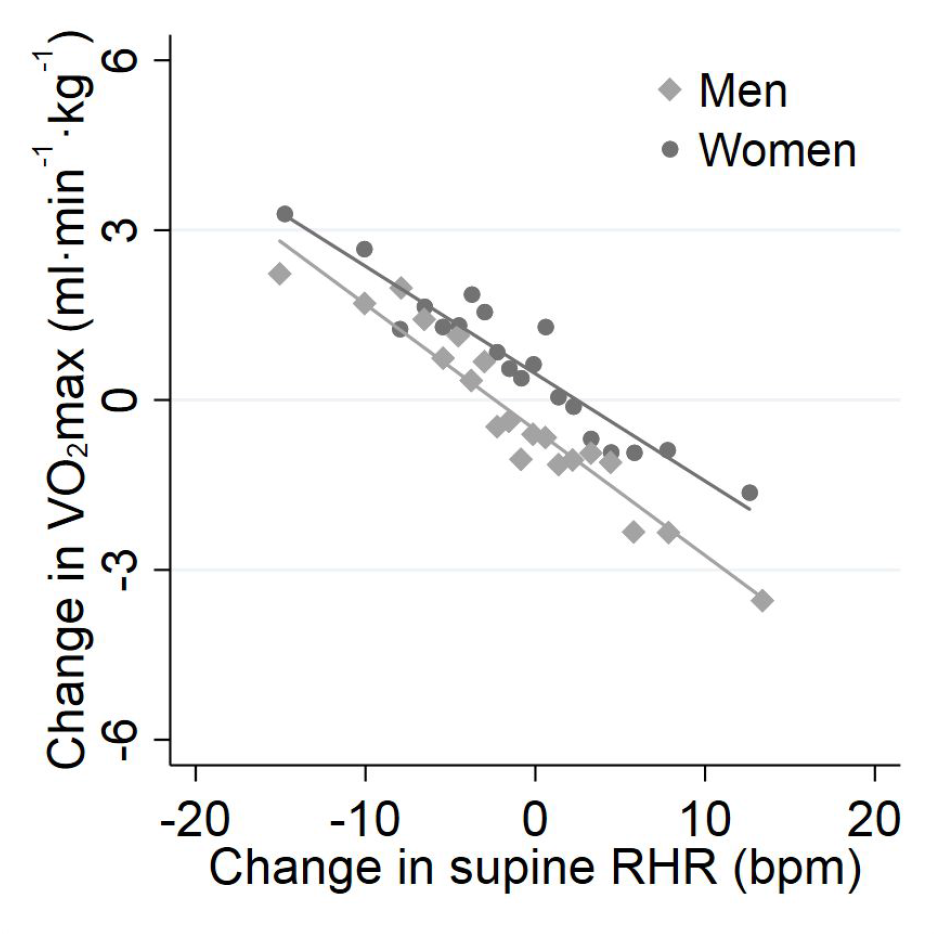
Association between 6-year change in supine resting heart rate and change in fitness, stratified by sex. Models were adjusted for adjusted for follow-up time and baseline values of age, sex, RHR, and VO_2_max. Longitudinal subsample, the Fenland Study (n=6,589). Each point represents 5% of the data in the binscatter plot.

## Discussion

We have documented the strong interrelationships between common measures of resting heart rate (RHR, when seated, lying supine, and during sleep) and investigated their relationship with cardiorespiratory fitness (CRF, estimated maximal oxygen consumption; VO_2_max) in a large population-based study of UK adults. Cross-sectional analyses showed a consistent inverse association between RHR and VO_2_max that was apparent across the different RHR measurement approaches and VO_2_max normalisation conventions (body mass, fat-free mass). A relatively small part of the association between RHR and VO_2_max was explained by adiposity but a much greater proportion was explained by the more readily modifiable behaviour physical activity. Concordantly in longitudinal analyses, within-person change in RHR was associated with within-person change in VO_2_max, similar in magnitude to the relationship that was observed cross-sectionally, thus supporting the notion that changes in factors determining CRF is paralleled by changes in RHR measures and hence RHR can be used as a biomarker for CRF.

This is the first study to examine these associations between multiple measures of RHR and CRF and interrogate them together with objective measures of adiposity and physical activity in a large cohort of men and women. We have previously shown that activity levels in this cohort are similar to those reported in national UK surveys (17,28), suggesting that CRF, as the capacity to undertake physical activity, may also be similar to national levels.

Previous detailed studies of the relationship between RHR and CRF have typically been small and focused on identifying electrophysiological predictors of exercise performance (29–32). There are, however, few large-scale studies that have explored this relationship with findings similar to those reported here. The Copenhagen Male Study found an inverse association between VO_2_max assessed submaximally in 1970 and supine RHR measured by 12-lead ECG about 15 years later in 2798 men (8); the RHR-to-CRF relationship (□ coefficient ≅ −0.19 ml O_2_ kg^-1^ beat^-1^) was shallower than values reported in the present study at around −0.30 ml O_2_ kg^-1^ beat^-1^. In the Danish Health Examination Survey, the relationship between seated RHR and VO_2_max was less pronounced (□ coefficient ≅ −0.12 ml O_2_ kg^-1^ beat^-1^) when assessed with maximal cycle ergometry in over 10 thousand men and women from 2007-8 (33,34). A weak prospective inverse association between RHR at baseline and VO_2_max at 23-year follow-up was reported in the Norwegian Nord-Trøndelag Health Study: −0.9 and −0.4 ml O_2_ kg^-1^ beat^-1^ in 807 men and 810 women, respectively (16). In the same study, within-person change in RHR between baseline and follow-up was inversely associated with VO_2_max at follow-up; within-person change in VO_2_max was not assessed. Among 56 thousand American patients referred by their physician for exercise testing and excluding those with known heart disease, the age-adjusted coefficient from meta-regression across RHR categories was −0.22 ml O_2_ kg^-1^ beat^-1^ (35), again similar to results reported in our present study, despite the difference in population sampling.

Our study is among the few to examine the influence of factors underpinning the RHR-to-CRF relationship. For all measures of RHR, we report significant inverse associations between RHR and CRF that are independent of age, sex, obesity and physical activity. The age, sex, BMI-, and physical activity-adjusted coefficient for seated RHR was about −0.13 ml O_2_ kg^-1^ beat^-1^. By comparison, a pooled cohort analysis of almost 50 thousand American and British individuals found a similarly adjusted RHR coefficient of about −0.17 ml O_2_ kg^-1^ beat^-1^ (36). However, physical activity was self-reported in those studies which may have inflated the value of the observed coefficients for RHR because of only partial adjustment for physical activity. In parallel, the Tromsø study compared seated RHR and CRF levels in 5017 men and 5607 women when stratified by self-reported activity levels (37), demonstrating significant inverse associations within sex and across physical activity strata. Together, these findings support the notion that RHR and habitual physical activity levels are intrinsically linked to exercise capacity. Thus, changes in CRF achieved through altered physical activity levels could be feasibly monitored with periodic RHR measurements. Additional work is needed to better elucidate the physical activity-CRF relationship, both cross-sectionally and longitudinally, so that the reliability of inferences from such an approach can be clarified.

It is well-recognised that RHR is associated with heart disease (18,38), diabetes (39,40), cancer (11,12), and all-cause, cardiovascular- and cancer-specific mortality (10,41) but the mechanisms underlying these are not fully understood. Knowing that both higher CRF and higher habitual physical activity levels are associated with lower incidence of such diseases and related mortality (5,42), the significant association of RHR with CRF and the degree to which that association is influenced by physical activity and body mass index offers an explanation for the association of RHR with these endpoints. In clinical practice, such as acute hospital settings, the use of RHR is primarily limited to the detection and management of bradycardia, tachycardia and related conditions. Our findings suggest that RHR may offer a relatively low-cost way of assessing an important risk factor for chronic disease among young-to-middle-aged adults and evaluating the effects of interventions targeting fitness in research, clinical practice and remote patient monitoring systems. We have facilitated this by documenting the interrelationships between common measures of RHR in the general population. However, for this to be directly clinically applicable, future work is needed to examine associations between RHR measures and their relation with such diseases in patient groups predisposed to reduced exercise capacity.

Our study has some limitations. We used heart response to a submaximal treadmill test to estimate rather than directly measure VO_2_max. Even though we have validated this approach (24), associations between RHR and CRF reported here may be influenced by residual error from the VO_2_max estimation process, which is largely dependent on reaching a percentage of age-predicted maximal HR. Additionally, VO_2_max estimated from HR response to submaximal exercise is unreliable in those taking medications such as beta-blockers (43). We therefore excluded participants on beta-blockers from analyses, as well as participants not passing the medical screening for treadmill testing and our results are unlikely to generalise to such individuals.

## Conclusions

We have addressed several methodological challenges associated with the estimation of CRF from RHR. In a population sample of UK adults we have shown that RHR is inversely associated with CRF across different RHR measurement approaches. Half of this association is explained by modifiable factors such as body size and habitual physical activity, suggesting that changes in RHR may be used to track changes in CRF over time. We further showed that within-person change in RHR was associated with within-person change in CRF. These findings position RHR as a feasible and responsive biomarker of CRF in the general population and in clinical care and research, facilitating personal goal setting, evaluation and remote patient monitoring.

## Data Availability

The data analysed during the current study are not publicly available because we have not obtained consent for public data sharing from the study participants.

## Acknowledgements

We are grateful to all Fenland Study participants who gave their time and effort. We also thank the functional teams of the MRC Epidemiology Unit at Cambridge (Field Epidemiology, Study Coordination, Data management and IT) for supporting this study.

## Author’s contribution

TG, JYJ, TL, and SB conceptualised the design of the present analysis. NF, SG, and SB are principal investigators and NW is chief investigator of the Fenland Study. KW and SH contributed to data collection and cleaning. TG, TL, and SB analysed the data. TG, JYJ, TL, IP, and SB drafted the manuscript. All authors provided critical feedback and helped shape the research, analysis, and approved the final version of the manuscript. TG, TL, and SB had full access to the data in the study and take responsibility for the integrity of the data and the accuracy of the data analysis. The corresponding author attests that all listed authors meet authorship criteria.

---

**Supplementary Figure 1.**
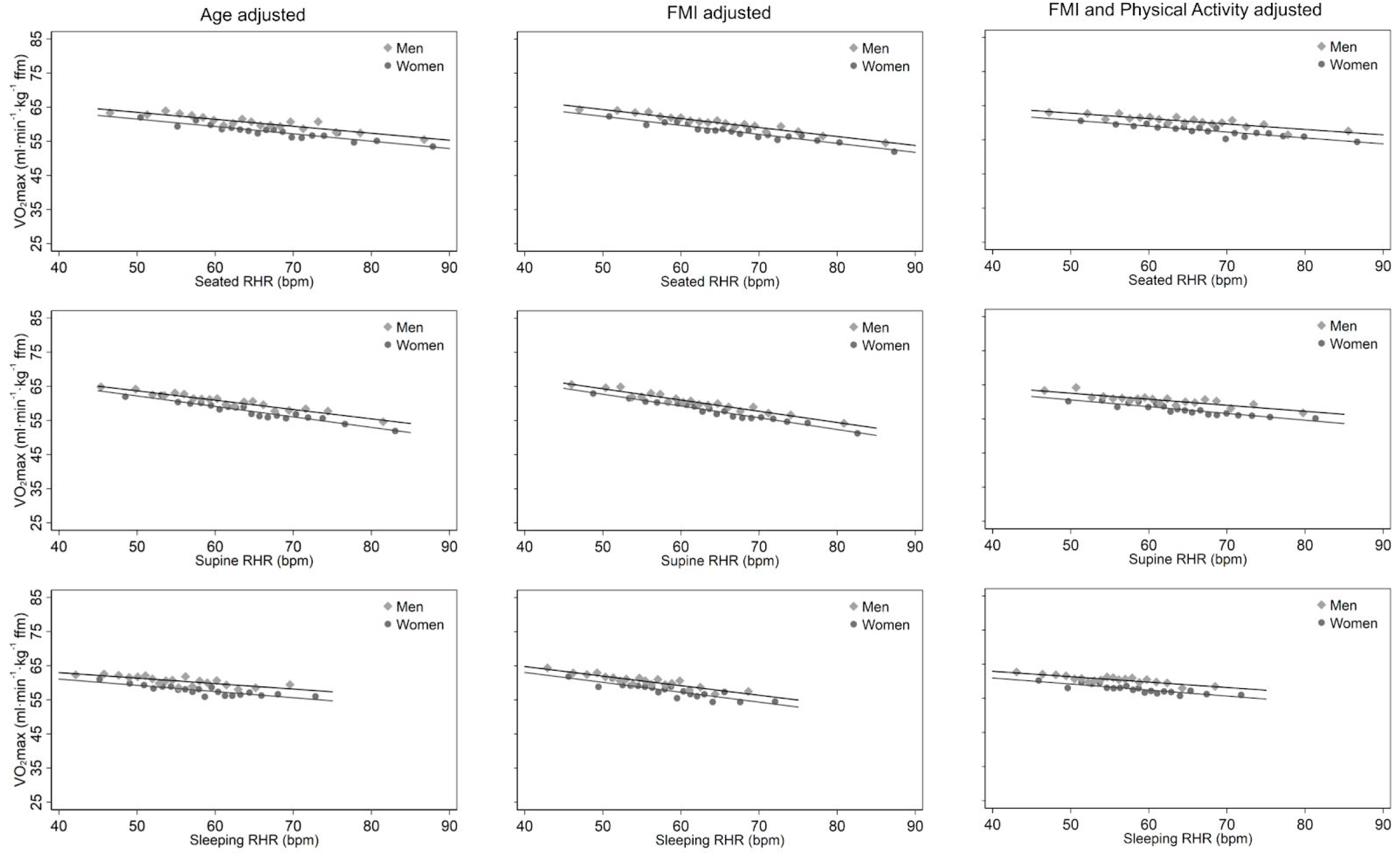
Associations between resting heart rate and maximal oxygen consumption expressed per kg fat-free mass, stratified by sex and adjusted for age (left column panels), age, ethnicity, smoking, alcohol consumption, and fat mass index (middle column panels), and further for physical activity energy expenditure (PAEE), moderate-vigorous PAEE (right column panels). Top: Sleeping resting heart rate. Middle: Supine resting heart rate. Bottom: Seated resting heart rate. The Fenland Study (n=10,865). Each point represents 5% of data in the binscatter plots.

**Supplementary Table 1.** Characteristics of complete cohort versus analytical sample in the Fenland study

|  | Fenland cohort (n=12,435) | Analytical sample (n=10,865) | P-value |
| --- | --- | --- | --- |
| Demographics |  |  |  |
| Sex (W/M) | 6,691 / 5,744 | 5772 / 5143 | 0.16 |
| Age (years) | 48.6 (7.5) | 48.2 (7.4) | 0.00 |
| Ethnicity |  |  | 0.50 |
| White | 11487 (92.4) | 10062 (92.6) |  |
| Non-White | 948 (7.6) | 803 (7.4) |  |
| Smoker Status |  |  | 0.12 |
| Never smoked | 6696 (53.9) | 5894 (54.3) |  |
| Ex-smoker | 4092 (32.9) | 3568 (32.8) |  |
| Current smoker | 1495 (12.0) | 1280 (11.8) |  |
| Alcohol |  |  | 0.80 |
| <1/week | 4196 (33.7) | 3536 (32.5) |  |
| 1-4/week | 6115 (49.2) | 5472 (50.4) |  |
| Almost daily | 1910 (15.4) | 1686 (15.5) |  |
| RHR |  |  |  |
| Seated (bpm) | 66.3 (9.8) | 66.1 (9.4) | 0.11 |
| Supine (bpm) | 63.3 (9.0) | 63.1 (8.6) | 0.08 |
| Sleeping (bpm) | 57.1 (7.0) | 56.9 (6.9) | 0.03 |
| Anthropometrics |  |  |  |
| Height (m) | 1.70 (0.09) | 1.71 (0.09) | 0.00 |
| Body mass (kg) | 78.2 (16.3) | 77.7 (15.7) | 0.02 |
| BMI (kg/m<sup>2</sup>) | 26.9 (4.8) | 26.6 (4.5) | 0.00 |
| FMI (kg/m<sup>2</sup>) | 9.1 (3.9) | 8.9 (3.6) | 0.00 |
| Percent body fat (%) | 33.0 (9.2) | 32.5 (9.0) | 0.00 |
| Physical Activity |  |  |  |
| PAEE kJ/day/kg | 53.6 (22.1) | 54.6 (22.0) | 0.00 |
| MVPA (min/day) | 102.2 (78.7) | 105.0 (78.6) | 0.01 |
| MVPA (kJ/day/kg) | 21.4 (17.5) | 22.0 (17.6) | 0.01 |
| Beta-blocker | 315 (2.5) | 0 (0) | 0.00 |

Values are means (standard deviation) or counts (%). RHR: Resting heart rate, bpm: beat per minute, BMI: Body mass index, FMI: Fat mass index, PAEE: Physical activity energy expenditure. MVPA: Moderate to vigorous physical activity.

**Supplementary Table 2.** Interrelationships between measures of resting heart rate. The Fenland Study (n=10,865).

| Equation | R<sup>2</sup> | RMSE (bpm) |
| --- | --- | --- |
| Supine RHR = 0.74 seated RHR + 14 | 0.65 | 5.1 |
| Supine RHR = 0.95 seated RHR | 0.65 | 5.4 |
| Sleeping HR = 0.58 supine RHR + 21 | 0.52 | 4.8 |
| Sleeping HR = 0.90 supine RHR | 0.52 | 5.5 |
| Sleeping HR = 0.47 seated RHR + 26 | 0.42 | 5.2 |
| Sleeping HR = 0.85 seated RHR | 0.42 | 6.3 |

R<sup>2</sup>: Coefficient of determination RMSE: Root mean squared error

**Supplementary Table 3.** Association between resting heart rate and maximal oxygen consumption expressed per kg fat-free mass. The Fenland Study.

|  |  | Seated RHR |  | Supine RHR |  | Sleeping RHR |  |
| --- | --- | --- | --- | --- | --- | --- | --- |
|  |  | Men | Women | Men | Women | Men | Women |
| Total sample |  |  |  |  |  |  |  |
| Model 1 |  | -0.21 (-0.25, -0.18) | -0.22 (-0.25, -0.18) | -0.28 (-0.32, -0.24) | -0.30 (-0.34, -0.26) | -0.17 (-0.22, -0.12) | -0.18 (-0.23, -0.13) |
| Model 2 |  | -0.21 (-0.25, -0.18) | -0.21 (-0.25, -0.18) | -0.27 (-0.31, -0.23) | -0.29 (-0.33, -0.26) | -0.20 (-0.25, -0.15) | -0.21 (-0.26, -0.16) |
| Model 3 |  | -0.27 (-0.30, -0.23) | -0.26 (-0.30, -0.22) | -0.33 (-0.37, -0.29) | -0.34 (-0.38, -0.30) | -0.28 (-0.33, -0.22) | -0.29 (-0.34, -0.23) |
| Model 4 |  | -0.19 (-0.23, -0.16) | -0.19 (-0.22, -0.15) | -0.22 (-0.25, -0.18) | -0.23 (-0.27, -0.19) | -0.18 (-0.23, -0.13) | -0.19 (-0.24, -0.14) |
| Model 5 |  | -0.17 (-0.20, -0.13) | -0.17 (-0.21, -0.14) | -0.18 (-0.22, -0.14) | -0.20 (-0.24, -0.16) | -0.16 (-0.20, -0.11) | -0.18 (-0.22, -0.13) |
| Age stratified |  |  |  |  |  |  |  |
| Model 2 | <50 years | -0.19 (-0.24, -0.13) | -0.21 (-0.27, -0.15) | -0.25 (-0.31, -0.20) | -0.26 (-0.32, -0.20) | -0.20 (-0.27, -0.12) | -0.19 (-0.26, -0.11) |
|  | 50-60 years | -0.21 (-0.26, -0.16) | -0.21 (-0.26, -0.15) | -0.25 (-0.31, -0.19) | -0.31 (-0.37, -0.24) | -0.16 (-0.24, -0.08) | -0.21 (-0.29, -0.13) |
|  | >60 years | -0.26 (-0.33, -0.18) | -0.24 (-0.33, -0.16) | -0.32 (-0.41, -0.24) | -0.34 (-0.44, -0.25) | -0.29 (-0.40, -0.17) | -0.26 (-0.38, -0.14) |
| Model 3 | <50 years | -0.24 (-0.29, -0.18) | -0.24 (-0.30, -0.19) | -0.32 (-0.37, -0.26) | -0.30 (-0.36, -0.24) | -0.28 (-0.36, -0.20) | -0.24 (-0.32, -0.16) |
|  | 50-60 years | -0.28 (-0.33, -0.22) | -0.27 (-0.32, -0.21) | -0.32 (-0.38, -0.26) | -0.36 (-0.42, -0.30) | -0.24 (-0.33, -0.16) | -0.30 (-0.38, -0.22) |
|  | >60 years | -0.30 (-0.37, -0.22) | -0.29 (-0.38, -0.21) | -0.37 (-0.45, -0.28) | -0.39 (-0.49, -0.30) | -0.32 (-0.44, -0.21) | -0.36 (-0.48, -0.24) |
| Model 5 | <50 years | -0.13 (-0.18, -0.08) | -0.13 (-0.19, -0.08) | -0.15 (-0.21, -0.09) | -0.13 (-0.19, -0.07) | -0.13 (-0.21, -0.06) | -0.11 (-0.19, -0.04) |
|  | 50-60 years | -0.18 (-0.23, -0.13) | -0.17 (-0.22, -0.11) | -0.17 (-0.23, -0.11) | -0.21 (-0.27, -0.15) | -0.14 (-0.22, -0.06) | -0.17 (-0.25, -0.09) |
|  | >60 years | -0.20 (-0.27, -0.12) | -0.25 (-0.34, -0.17) | -0.22 (-0.31, -0.14) | -0.32 (-0.41, -0.22) | -0.21 (-0.32, -0.10) | -0.31 (-0.43, -0.19) |
| BMI stratified |  |  |  |  |  |  |  |
| Model 3 | <25 kg/m<sup>2</sup> | -0.33 (-0.39, -0.27) | -0.29 (-0.34, -0.24) | -0.37 (-0.44, -0.31) | -0.38 (-0.44, -0.33) | -0.32 (-0.41, -0.23) | -0.36 (-0.43, -0.29) |
|  | 25-29.99 kg/m<sup>2</sup> | -0.24 (-0.29, -0.19) | -0.27 (-0.34, -0.19) | -0.33 (-0.38, -0.27) | -0.32 (-0.39, -0.24) | -0.27 (-0.34, -0.19) | -0.23 (-0.32, -0.13) |
|  | >30 kg/m<sup>2</sup> | -0.24 (-0.31, -0.16) | -0.19 (-0.28, -0.11) | -0.29 (-0.38, -0.21) | -0.31 (-0.39, -0.22) | -0.24 (-0.35, -0.12) | -0.21 (-0.33, -0.10) |
| Model 5 | <25 kg/m<sup>2</sup> | -0.20 (-0.26, -0.14) | -0.17 (-0.22, -0.12) | -0.19 (-0.26, -0.12) | -0.21 (-0.27, -0.16) | -0.17 (-0.25, -0.08) | -0.23 (-0.30, -0.16) |
|  | 25-29.99 kg/m<sup>2</sup> | -0.15 (-0.20, -0.11) | -0.19 (-0.26, -0.12) | -0.19 (-0.25, -0.14) | -0.20 (-0.27, -0.12) | -0.18 (-0.25, -0.11) | -0.13 (-0.22, -0.04) |
|  | >30 kg/m<sup>2</sup> | -0.14 (-0.21, -0.06) | -0.14 (-0.22, -0.06) | -0.16 (-0.24, -0.07) | -0.21 (-0.30, -0.12) | -0.11 (-0.22, -0.00) | -0.14 (-0.25, -0.03) |
| PAEE stratified |  |  |  |  |  |  |  |
| Model 5 | <40 kJ/day/kg | -0.16 (-0.23, -0.10) | -0.24 (-0.30, -0.18) | -0.23 (-0.31, -0.15) | -0.30 (-0.36, -0.23) | -0.13 (-0.24, -0.03) | -0.20 (-0.29, -0.11) |
|  | 40-60 kJ/day/kg | -0.20 (-0.25, -0.14) | -0.15 (-0.20, -0.09) | -0.22 (-0.28, -0.15) | -0.17 (-0.23, -0.11) | -0.17 (-0.25, -0.09) | -0.17 (-0.25, -0.10) |
|  | >60 kJ/day/kg | -0.13 (-0.19, -0.08) | -0.10 (-0.17, -0.03) | -0.11 (-0.17, -0.05) | -0.11 (-0.18, -0.03) | -0.16 (-0.23, -0.09) | -0.16 (-0.25, -0.07) |

Reported values are $\beta$ coefficients (95% confidence interval)
Model 1: Age adjusted
Model 2: Model 1 + ethnicity, smoking and alcohol adjusted
Model 3: Model 2 + fat mass index adjusted
Model 4: Model 3 + physical activity energy expenditure (PAEE) adjusted
Model 5: Model 4 + moderate-vigorous intensity PAEE adjusted

## References

1. Katzmarzyk PT, Church TS, Janssen I, Ross R, Blair SN. Metabolic syndrome, obesity, and mortality: impact of cardiorespiratory fitness. Diabetes Care. 2005;28(2):391–397.

2. Wei M, Gibbons LW, Mitchell TL, Kampert JB, Lee CD, Blair SN. The association between cardiorespiratory fitness and impaired fasting glucose and type 2 diabetes mellitus in men. Ann Intern Med. 1999;130(2):89–96.

3. Schmid D, Leitzmann MF. Cardiorespiratory fitness as predictor of cancer mortality: a systematic review and meta-analysis. Ann Oncol. 2015 Feb;26(2):272–8.

4. Blair SN, Kampert JB, Kohl HW, Barlow CE, Macera CA, Paffenbarger RS, et al. Influences of cardiorespiratory fitness and other precursors on cardiovascular disease and all-cause mortality in men and women. Jama. 1996;276(3):205–210.

5. Kodama S, Saito K, Tanaka S, Maki M, Yachi Y, Asumi M, et al. Cardiorespiratory fitness as a quantitative predictor of all-cause mortality and cardiovascular events in healthy men and women: a meta-analysis. JAMA. 2009 May 20;301(19):2024–35.

6. Jensen MT, Marott JL, Allin KH, Nordestgaard BG, Jensen GB. Resting heart rate is associated with cardiovascular and all-cause mortality after adjusting for inflammatory markers: the Copenhagen City Heart Study. Eur J Prev Cardiol. 2012 Feb;19(1):102–8.

7. Jensen MT, Marott JL, Jensen GB. Elevated resting heart rate is associated with greater risk of cardiovascular and all-cause mortality in current and former smokers. Int J Cardiol. 2011 Sep 1;151(2):148–54.

8. Jensen MT, Suadicani P, Hein HO, Gyntelberg F. Elevated resting heart rate, physical fitness and all-cause mortality: a 16-year follow-up in the Copenhagen Male Study. Heart. 2013 Jun 15;99(12):882–7.

9. Cooney MT, Vartiainen E, Laakitainen T, Juolevi A, Dudina A, Graham IM. Elevated resting heart rate is an independent risk factor for cardiovascular disease in healthy men and women. Am Heart J. 2010 Apr;159(4):612-619.e3.

10. Zhang D, Wang W, Li F. Association between resting heart rate and coronary artery disease, stroke, sudden death and noncardiovascular diseases: a meta-analysis. CMAJ [Internet]. 2016 Jan 1 [cited 2020 Mar 26]; Available from: https://www.cmaj.ca/content/early/2016/08/22/cmaj.160050

11. Lee DH, Park S, Lim SM, Lee MK, Giovannucci EL, Kim JH, et al. Resting heart rate as a prognostic factor for mortality in patients with breast cancer. Breast Cancer Res Treat. 2016 Sep;159(2):375–84.

12. Seviiri M, Lynch BM, Hodge AM, Yang Y, Liew D, English DR, et al. Resting heart rate, temporal changes in resting heart rate, and overall and cause-specific mortality. Heart Br Card Soc. 2018;104(13):1076–85.

13. Turjanmaa V, Kalli S, Sydänmaa M, Uusitalo A. Short-term variability of systolic blood pressure and heart rate in normotensive subjects. Clin Physiol. 1990 Jul;10(4):389–401.

14. Oja P. Dose response between total volume of physical activity and health and fitness. Med Sci Sports Exerc. 2001 Jun;33(Supplement):S428–37.

15. LaMonte MJ, Blair SN. Physical activity, cardiorespiratory fitness, and adiposity: contributions to disease risk: Curr Opin Clin Nutr Metab Care. 2006 Sep;9(5):540–6.

16. Nauman J, Aspenes ST, Nilsen TIL, Vatten LJ, Wisløff U. A Prospective Population Study of Resting Heart Rate and Peak Oxygen Uptake (the HUNT Study, Norway). Kiechl S, editor. PLoS ONE. 2012 Sep 18;7(9):e45021.

17. Lindsay T, Westgate K, Wijndaele K, Hollidge S, Kerrison N, Forouhi N, et al. Descriptive epidemiology of physical activity energy expenditure in UK adults (The Fenland study). Int J Behav Nutr Phys Act. 2019 09;16(1):126.

18. Böhm M, Reil J-C, Deedwania P, Kim JB, Borer JS. Resting heart rate: risk indicator and emerging risk factor in cardiovascular disease. Am J Med. 2015 Mar;128(3):219–28.

19. Brage S, Brage N, Franks PW, Ekelund U, Wareham NJ. Reliability and validity of the combined heart rate and movement sensor Actiheart. Eur J Clin Nutr. 2005 Apr;59(4):561–70.

20. Brage S, Ekelund U, Brage N, Hennings MA, Froberg K, Franks PW, et al. Hierarchy of individual calibration levels for heart rate and accelerometry to measure physical activity. J Appl Physiol. 2007 Aug 1;103(2):682–92.

21. Tanaka H, Monahan KD, Seals DR. Age-predicted maximal heart rate revisited. J Am Coll Cardiol. 2001 Jan;37(1):153–6.

22. Henry CJK. Basal metabolic rate studies in humans: measurement and development of new equations. Public Health Nutr. 2005 Oct;8(7A):1133–52.

23. Consolazio C, Johnson R, Pecora L. Physiological measurements of metabolic functions in man. New York: McGraw Hill; 1963.

24. Gonzales TI, Westgate K, Hollidge S, Lindsay T, Jeon J, Brage S. Estimating maximal oxygen consumption from heart rate response to submaximal ramped treadmill test. medRxiv. 2020 Feb 20;2020.02.18.20024489.

25. Mazess RB, Barden HS, Bisek JP, Hanson J. Dual-energy x-ray absorptiometry for total-body and regional bone-mineral and soft-tissue composition. Am J Clin Nutr. 1990 Jun;51(6):1106–12.

26. Stegle O, Fallert SV, MacKay DJC, Brage S. Gaussian process robust regression for noisy heart rate data. IEEE Trans Biomed Eng. 2008 Sep;55(9):2143–51.

27. Brage S, Brage N, Franks PW, Ekelund U, Wong M-Y, Andersen LB, et al. Branched equation modeling of simultaneous accelerometry and heart rate monitoring improves estimate of directly measured physical activity energy expenditure. J Appl Physiol. 2004 Jan 1;96(1):343–51.

28. Brage S, Lindsay T, Venables M, Wijndaele K, Westgate K, Collins D, et al. Descriptive epidemiology of energy expenditure in the UK: findings from the National Diet and Nutrition Survey 2008–15. Int J Epidemiol. 2020;0(0):15.

29. Grant CC, Murray C, Janse van Rensburg DC, Fletcher L. A comparison between heart rate and heart rate variability as indicators of cardiac health and fitness. Front Physiol [Internet]. 2013 [cited 2020 May 19];4. Available from: http://journal.frontiersin.org/article/10.3389/fphys.2013.00337/abstract

30. Billman GE, Cagnoli KL, Csepe T, Li N, Wright P, Mohler PJ, et al. Exercise training-induced bradycardia: evidence for enhanced parasympathetic regulation without changes in intrinsic sinoatrial node function. J Appl Physiol. 2015 Mar 6;118(11):1344–55.

31. Sartor F, Bonato M, Papini G, Bosio A, Mohammed RA, Bonomi AG, et al. A 45-second self-test for cardiorespiratory fitness: Heart rate-based estimation in healthy individuals. Fukumoto Y, editor. PLOS ONE. 2016 Dec 13;11(12):e0168154.

32. Plaza-Florido A, Migueles JH, Mora-Gonzalez J, Molina-Garcia P, Rodriguez-Ayllon M, Cadenas-Sanchez C, et al. Heart Rate Is a Better Predictor of Cardiorespiratory Fitness Than Heart Rate Variability in Overweight/Obese Children: The ActiveBrains Project. Front Physiol. 2019 May 7;10:510.

33. Eriksen L, Grønbaek M, Helge JW, Tolstrup JS. Cardiorespiratory fitness in 16 025 adults aged 18-91 years and associations with physical activity and sitting time. Scand J Med Sci Sports. 2016 Dec;26(12):1435–43.

34. Helge J. personal communication. 2020.

35. Aladin AI, Whelton SP, Al-Mallah MH, Blaha MJ, Keteyian SJ, Juraschek SP, et al. Relation of Resting Heart Rate to Risk for All-Cause Mortality by Gender After considering Exercise Capacity (the Henry Ford Exercise Testing Project). Am J Cardiol. 2014 Dec;114(11):1701–6.

36. Jurca R, Jackson AS, LaMonte MJ, Morrow JR, Blair SN, Wareham NJ, et al. Assessing Cardiorespiratory Fitness Without Performing Exercise Testing. Am J Prev Med. 2005 Oct;29(3):185–93.

37. Emaus A, Degerstrøm J, Wilsgaard T, Hansen BH, Dieli-Conwright CM, Furberg A-S, et al. Does a variation in self-reported physical activity reflect variation in objectively measured physical activity, resting heart rate, and physical fitness? Results from the Tromsø study. Scand J Public Health. 2010 Nov;38(5_suppl):105–18.

38. Böhm M, Schumacher H, Teo KK, Lonn EM, Mahfoud F, Ukena C, et al. Resting heart rate and cardiovascular outcomes in diabetic and non-diabetic individuals at high cardiovascular risk analysis from the ONTARGET/TRANSCEND trials. Eur Heart J. 2020 Jan 7;41(2):231–8.

39. Lee DH, Rezende LFM de, Hu FB, Jeon JY, Giovannucci EL. Resting heart rate and risk of type 2 diabetes: A prospective cohort study and meta-analysis. Diabetes Metab Res Rev. 2019;35(2):e3095.

40. Yang HI, Kim HC, Jeon JY. The association of resting heart rate with diabetes, hypertension, and metabolic syndrome in the Korean adult population: The fifth Korea National Health and Nutrition Examination Survey. Clin Chim Acta Int J Clin Chem. 2016 Apr 1;455:195–200.

41. Aune D, Sen A, ó’Hartaigh B, Janszky I, Romundstad PR, Tonstad S, et al. Resting heart rate and the risk of cardiovascular disease, total cancer, and all-cause mortality - A systematic review and dose-response meta-analysis of prospective studies. Nutr Metab Cardiovasc Dis NMCD. 2017 Jun;27(6):504–17.

42. Moore SC, Lee I-M, Weiderpass E, Campbell PT, Sampson JN, Kitahara CM, et al. Association of leisure-time physical activity with risk of 26 types of cancer in 1.44 million adults. JAMA Intern Med. 2016 01;176(6):816–25.

43. Wonisch M, Hofmann P, Fruhwald FM, Kraxner W, H??dl R, Pokan R, et al. Influence of beta-blocker use on percentage of target heart rate exercise prescription: Eur J Cardiovasc Prev Rehabil. 2003 Aug;10(4):296–301.

